# Understanding the implementation of a smoking cessation intervention for people experiencing homelessness: A process evaluation of the SCeTCH trial

**DOI:** 10.1101/2025.05.06.25327053

**Authors:** Rachel Brown, Allison Ford, Laura Reid, Lauren McMillan, Kirstie Soar, Francesca Pesola, Caitlin Notley, Emma Ward, Bethany Gardner, Anna Varley, Charlotte Mair, Jessica Lennon, Janine Brierley, Amy Edwards, Danielle Mitchell, Debbie Robson, Peter Hajek, Allan Tyler, Steve Parrott, Jinshuo Li, Linda Bauld, Sharon Cox

## Abstract

**Introduction:** Cigarette smoking is higher in people experiencing homelessness. The Smoking Cessation Trial in Centres for Homelessness (SCeTCH) was a two-armed cluster randomised controlled trial in 32 services for homelessness across Great Britain. We compared the provision of an e-cigarette (EC) starter kit with usual care (UC). Findings from the process evaluation, focusing on intervention implementation from the perspectives of staff at EC centres and researchers are reported.

**Methods:** The process evaluation involved qualitative interviews with staff (N=16) and participants (N=31), observations of intervention delivery (N=32), fidelity checklists (N=25) (at EC sites), training evaluations (N=160) and data collected within the main trial (at both EC and UC sites). Findings here are from EC sites.

**Results:** Findings suggest that it is feasible to deliver a smoking cessation intervention in these services. The EC intervention was largely acceptable to staff and the training we delivered was highly rated. Individual intervention components were delivered with high fidelity, with no differences by centre size or location, suggesting high transferability.

**Conclusion:** Challenges to future delivery of similar interventions may stem from lack of wider staff team buy-in and support, as well as challenges to time available when faced with more immediate service user needs.

## Introduction

Rates of cigarette smoking are significantly higher among people experiencing homelessness than the population who are housed, with UK data showing smoking rates of 11.9% among the adult population as a whole^1^, compared with estimates between 76-85% among those who are homeless^2,3^. Higher tobacco use contributes to already poor health outcomes including significantly higher rates of respiratory and cardiovascular disease and illness than those who are securely housed^4^. People who are experiencing homelessness are more likely to engage in potentially-higher risk smoking behaviours such as sharing cigarettes, smoking discarded cigarettes and smoking unfiltered tobacco^5^ which may exacerbate the risk of respiratory illnesses.

Motivation to quit and past-year attempts are no different compared with people who are not experiencing homelessness^6,7^. In the UK, despite access to local stop smoking services (SSS) and free cessation medications, use of the SSS is low among people experiencing homelessness and quit success is short-lived, often lasting no more than 24 hours^5,8^. To date, there is little high-certainty evidence for which smoking cessation approaches are most effective for those who experience homelessness, but some evidence suggests that higher intensity interventions may be more effective^9^. In addition, qualitative studies have shown that perceived ability to quit is much higher when people feel well supported in making the change^6^, and where other people who are ex-smokers or trying to quit are present to model and normalise quitting^10^.

However, accessing support for quitting in an environment which also helps address other health and social needs presents challenges. Around half of services supporting people who are homeless report screening their service users for smoking and a similar number have made a referral to their local smoking cessation service. However, referrals are rarely taken up, and only 12% of services having established links with their local stop smoking service^8^. The frequency and volume of smoking in homeless centres is a commonly cited barrier to quitting, with smoking being cited a key part of the culture and social interactions with both service users and staff^7,11^. Smoking rates among staff working in the homeless sector are also around twice as high as the general population, and smoking with service users as a means of casual interaction is common^8^.

In the UK, e-cigarettes are the most used aid to quit smoking^12^ and pooled evidence from trial and observational studies show they are an effective quit aid^13, 14^. There are also some signs of promise that these may help people accessing homeless services to reduce or quit smoking. Previously, we explored the feasibility of providing a free e-cigarette starter pack to people accessing homeless centres, along with information for centre staff on e-cigarette use and quitting^7,8,15^. This study demonstrated the feasibility of a full trial, leading to the development of the Smoking Cessation Trial in Centres for Homelessness (SCeTCH). The SCeTCH study was a multi-site two arm cluster randomised controlled trial taking place in 6 regions across Great Britain^16^ (ISRCTN registry ID:18566874). A summary is provided in Figure 1 below.

The trial included a mixed method process evaluation exploring the hypothesised mechanisms of the intervention and contextual influences on implementation (see https://osf.io/yhmk9/), including the role of centre smoking culture and norms.

Understanding how the trial and intervention were implemented is important for future research and implementation of similar interventions within similar services, and so is the aim of the current paper. Here, we draw on the RE-AIM Framework^17^ (Glasgow et al. 1999) which has been developed to improve implementation of public health interventions by assessing five dimensions: Reach, Effectiveness, Adoption,

Implementation, and Maintenance, and frame our data from various study sources around this. Re-aim has been used effectively in assessing health interventions in community settings^18,19^ and in mapping process evaluation data on intervention implementation^20^.

## Methods

The SCeTCH trial included an embedded, mixed method process evaluation involving qualitative interviews with centre staff and participants, observations of intervention delivery, fidelity checklists, training evaluations and data embedded in main trial data collection (case report forms completed at baseline, 4, 12 and 24 weeks follow-up). Ethical approval for the trial was obtained from London South Bank University ethics committee (ETH2021-0176/ETH2122-0130 (details of how consent was given is provided below).

Three research questions underpinned the process evaluation:

- RQ1: How is the EC intervention implemented and how does organisational and geographic context influence implementation?
- RQ2: What are the mechanisms through which the delivered intervention activities and participant interactions produce change in smoking behaviour?
- RQ3: If the intervention is effective and cost-effective, what are the facilitators and barriers to roll out across Great Britain?

This paper focuses on RQ1. Full details of trial methods, registration and ethical approvals can be found at https://doi.org/10.1111/add.1585116. Methods and findings are summarised in Supplement 1, (reproduced from Ford et al. 2025 pre-print^21^).

### Sampling and recruitment

Thirty-two centres providing support to people experiencing homelessness were recruited across six regions, comprising: Wales and South West England (N=6), Scotland (n=6), London (N=7), East of England (N=7), Southeast England (N=6). The initial aim was to recruit support centres providing open-access day services without residential status, mirroring the feasibility study^7^. However, mapping of available services identified a lack of such services in regions outside of London, with the decision to extend recruitment to support centres providing accommodation alongside keyworker support, advice and guidance. Centres providing emergency accommodation only were excluded.

Within the 32 participating sites, 477 service users were recruited for the full trial (239 in the EC arm). A sub-sample of 8 sites (2 in Wales, 2 in Scotland and 4 in England) were identified for the process evaluation, representing a range of centre size, urban/rural location and range of support services offered. Participant demographics are reported in full in Ford et al. (2025 – preprint^21^).

### Data collection and analysis

#### Staff training

220 staff across the sites took part in training delivered by research staff (109 in EC centres). Training took between 2-3 hours and staff were invited to complete a one-page evaluation at the end, covering opinions of the length of the training, the content and structure, the session timing and the opportunity for them ask questions following the training. A total of 164 training evaluation forms were returned and analysed to provide descriptive statistics. Of these, 79 were completed in the EC arm and are reported here.

#### Fidelity checklists

Researchers completed observations of EC intervention delivery by centre staff, firstly at the initial delivery of the intervention and then again at weeks 3-4 at follow-up meetings between staff and service users. This involved a checklist (Appendix A) to assess quality and consistency of delivery and researchers were also able to add narratives to checklists to provide details of relevance on context, centre issues on the day of observation, participant responses etc. A total of 24 checklists were completed. These checklists were analysed using tick-box metrics to provide descriptive statistics. Narrative accounts were analysed thematically.

#### Ethnographic observations

Research staff completed a structured observation sheet (Appendix B) at regular intervals on visits to support centres to capture staffing levels at the time of data collection, participant numbers, services being offered, site details e.g., which areas smoking is permitted and data on the visible promotion of the study. A total of 32 observation forms were completed with narrative accounts and were analysed thematically (16 at EC sites, reported on here).

#### Qualitative interviews

A total of 31 service users and 16 staff at EC sites took part in semi-structured interviews. Written consent was obtained for interview and audio recording (separate to initial participant consent for the main trial). Consideration was given to ensuring that people had capacity to consent at the time of interview, with alternative appointment dates made.

Service user interviews took place in centres in quiet spaces to ensure privacy, however centre staff were aware of interviews taking place for researcher and participant safety.

Topic guides were developed to explore hypothesised methods of change and are reported elsewhere^21^. These were then examined by a Patient and Public Involvement (PPI) group providing support to the trial. Service user interviews took place between 12 and 24 week follow ups and included discussion of smoking, vaping and other health behaviours, experience of taking part in the trial in their specific centre, barriers and facilitators of changes to smoking behaviour.

Staff interviews were completed by telephone by researchers not known to them (from other regions), this was planned to avoid any social desirability effects around reporting positively to those who had delivered the training. The interviews included discussion around the barriers and facilitators to the intervention at their centre, perceived acceptability of both the intervention and the trial methods and opinions on potential options for sustainable future delivery of such interventions. Staff interviews were completed between weeks 4-8 of intervention delivery.

Transcripts of qualitative interviews were analysed thematically^22^ (Braun and Clarke, 2019) combining inductive and deductive approaches to explore study research questions and emerging themes. We used the RE-AIM framework^17^ as a guide for categorising themes in order to understand how each theme was important for specific implementation development activities going forward. Separate coding frames were initially developed for service user and staff interviews from open reading of a sub-sample of transcripts. Researchers (EW, LM, JL and AF) then independently tested these against a further sub-sample, before refinements were made through group discussion. Full data sets were then coded using NVivo 12.

## Results

Findings are presented drawing on the RE-AIM framework^17^ developed to improve implementation of public health interventions by assessing five dimensions: Reach, Effectiveness, Adoption, Implementation, and Maintenance. Table 1 (adapted from Welch et al. 2020^20^) summarises the structure of this section, including dimensions of the framework reported here and data sources. Findings focus on intervention delivery in the EC arm of the trial, in line with the emphasis on understanding intervention implementation.

**Table 1:** PE data sources and findings mapped onto the domains of the Re-aim Framework.

| Criteria | Data source |
| --- | --- |
| **Reach** |  |
| **Definition:** Summary of participant recruitment. | *Full details of participant recruitment are reported elsewhere<sup>23</sup>* |
| **Effectiveness** |  |
| **Definition:** the impact of the intervention on primary (sustained smoking abstinence) and secondary (short-term abstinence and smoking reduction) outcomes, as well as other secondary outcome measures collected. | *Main outcomes are reported elsewhere<sup>23</sup>.* |
| **Adoption—setting level** |  |
| **Definition:** the number of support centres that committed to participation in the study compared to those who were approached and did not participate | *The number of centres approached and the number of centres that took part* |
| **Adoption—individual staff level** |  |
| **Definition:** the number and type of centre staff that committed to participation in the study compared to total centre staff | *Summaries taken from the staff training evaluation forms*<br>*Staff interviews on the reflections on whether the training was adequate for delivery as intended and any other barriers* |
| **Implementation** |  |
| **Definition:** the extent to which the intervention was implemented as intended, as well as its consistency across centres and over the intervention period. |  |
| Delivery (e.g., fidelity) | *Fidelity checklists and researchers' observations of intervention delivery - as intended – and notes of any adaptations*<br>*Staff interview data on the challenges of the delivery as intended.* |
| Cost of intervention—time | *Reported elsewhere in economic evaluation and not reported here.* |
| Cost of intervention—money | *Reported elsewhere in economic evaluation and not reported here.* |
| Consistency of implementation across staff/time/settings/subgroups | *Differences in intervention delivery by: centre type, nation, throughput of service users, staffing, as recorded in researcher observations*<br>*Staff interview data on barriers and facilitators for delivering the intervention.* |
| **Maintenance—individual level** | Reported elsewhere<sup>21,23</sup> |
| **Definition:** Primary and secondary outcomes at 24-week follow up |  |
| **Maintenance—setting level** |  |
| **Definition:** the extent to which behaviour changes were implemented in participating centres after the intervention delivery period. | • *Evidence of ongoing activity in centres, including any changes in smoking locations and centre culture, from staff interviews and researcher observations.*<br>• *Any ongoing smoking cessation conversations or new services within centres including any links with local cessation services.* |
| Consideration of perceived alignment to organisation mission | • *Acceptability as captured in:*<br>• *Staff interviews on perceived ease of delivering the intervention and 'fit' with centres practices<sup>21</sup>* |

### Reach

In participating centres, a target was set of 15 eligible service users taking part in the trial per site, with actual recruitment ranging from 12-17. There were challenges in recruiting at UC sites where it was perceived that the offering was more limited than in EC sites, as well as in residential settings where residents can spend large amounts of time in their own rooms rather than in shared areas. Recruitment was also challenging in smaller centres with fewer service users overall who met the criteria for inclusion. In some areas, such as many local authorities in Wales, smaller residential centres are more typical due to lower population numbers and the tendency for people experiencing homelessness to migrate to urban centres. Despite these challenges, overall participant recruitment close to the target of 480 was achieved (N=477) and achieved largely within the allotted timeframe.

There were multiple approaches to service user recruitment. Initially, on first site visit, prior to randomisation into trial arms, expression of interest forms were displayed in centres, and staff were asked to explain that a research study was being conducted and to engage participants. Research staff also spent time on site in communal areas (e.g., canteens and activity rooms), engaging service users in conversation and eliciting interest. We found that becoming a ‘known face’ on site and building trust with service users was effective for supporting recruitment and was aided by how engaged staff were in promoting the study, e.g. introducing the researcher to people. Observations noted differences in centres in the perceived quality of relationships between staff and service users, in terms of friendliness of conversations, greetings of people entering and time spent together. In the majority, it was felt that relationships were friendly and open, and staff were highly supportive, even in centres which were often short-staffed or impacted by high turnover of staff and volunteers. In a smaller number of centres, these relationships were seen as less positive with a notable sense of separation between staff and service users. One researcher observed that participant recruitment was more difficult where staff/service user relationships appeared to be more strained.

### Adoption at centre level

A total of 183 centres were approached to participate in the study. Forty-seven agreed to a meeting with research staff, 26 services declined to take part and the remainder did not respond to contact. Of those who declined, reasons given were varied and included lack of staff capacity, concerns over ongoing service funding, being allocated to usual care and willingness to address smoking due to other behaviours being prioritised. At the time recruitment commenced, services were still impacted by the Covid-19 pandemic, with some still closed to non-residents, some partially open or in the process of re-opening and many stating that their staffing and volunteer numbers had been adversely affected and not yet recovered to previous levels.

Initial approaches to centres were made by area leads in each of the regions. Where possible this involved an in-person visit to the centre to speak to the manager and other available staff. The study was outlined, including both trial arms, the randomisation process and intended researcher input in the centre. From these visits the target of 32 participating sites was met. These were randomised (16 EC and 16 UC arms), at which point one site withdrew due to being allocated to UC. A replacement site meeting the inclusion criteria was recruited. The in-person approach was effective in centre engagement, with opportunities to answer questions and assure staff that the research team would work flexibly and adapt the timing/length of visits etc. to centre needs.

### Adoption at individual staff level

220 members of staff across all sites received an introductory session (109 in EC sites, including the VBA+ training package^24^), delivered in-person by members of the research team in sessions lasting between 2-3 hours. The session was developed to be delivered in three hours, however many centres were unable to free their staff for this length of time and requested that it be shortened. Shortening generally involved reducing the amount of background information on smoking harms but retaining the content on e-cigarettes (at EC sites) and how to use them to retain the core components of the intervention. 79 staff at EC sites completed a training evaluation form at the end of the session, with over 90% of respondents either strongly agreeing or agreeing with seven key statements on training content (see Table 2), suggesting that the training constituted effective preparation for intervention delivery.

**Table 2:**
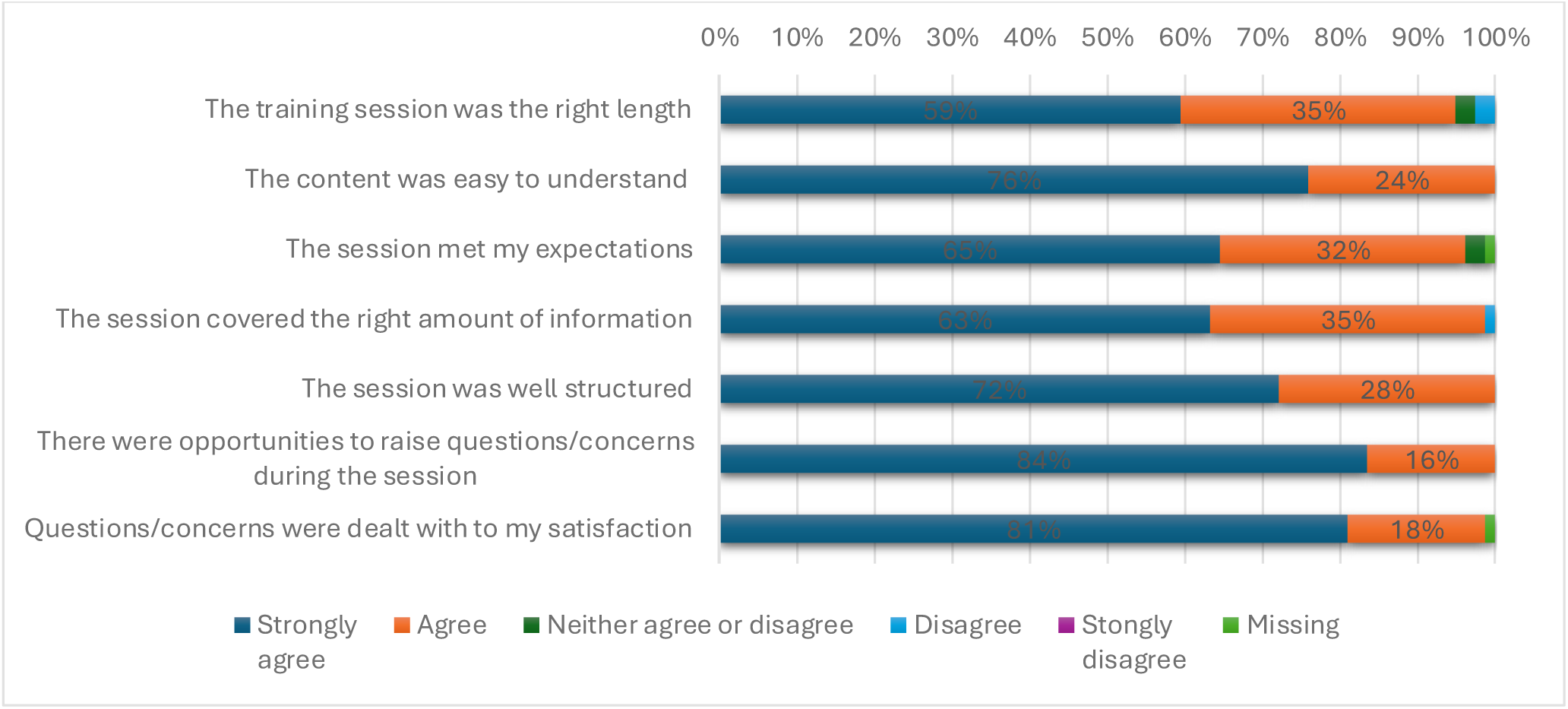
Training feedback for staff in the EC arm.

Open-ended responses were invited on what staff found most useful about the training. Responses included: increased understanding of smoking harms compared to e-cigarette use; understanding more about smoking harms for people experiencing homelessness; being able to challenge media stories on e-cigarette harms. These themes were also identified in interviews, with staff reporting the training was effective in alleviating their own pre-existing concerns around e-cigarettes:

> Personally before the training I was like, oh no, you know, I don’t think they’re good for you, there’s not been enough research, you know, I don’t know much about them. But after the training I realised actually they’re quite a good thing. (Female, Manager)

Some found it useful to be able to address participants concerns about using the devices, including those related to common media narratives at the time of the study, for example:

> …a couple of folk came and spoke to me about it, oh they didn’t want to do it because of [popcorn lung]…and I explained where the popcorn lung came from…it’s got nothing to do with e-cigarettes…it was great that I was able to explain that to them, after being explained to me. (Female, Support Worker)

While all staff that attended the EC training were theoretically able to then offer the intervention to service users, researcher observations illustrated that a much smaller number often went on to deliver it, with this usually being led by 1 or 2 key people in the centre. In many cases, these were staff who had a specific wellbeing remit in their job title/role and/or were users of e-cigarettes themselves and potentially more interested in the intervention. Other staff who had attended the training would then refer their service users to this person.

### Implementation fidelity

Fidelity checklists were completed at two time points, with two per EC centre at the point of initial intervention delivery (N=16) and a further follow-up ‘check in’ between participants and staff during the 4-week intervention period (N=8). For initial intervention delivery, fidelity checklists were designed around the core intervention components outlined in the training. Results are displayed in Table 3:

**Table 3.** Summary of completed fidelity checklists.

| *Component* | *Discussed*<br>*(N out of 16)* | *Not*<br>*discussed (N*<br>*out of 16)* |
| --- | --- | --- |
| *Before visit, checked EC is fully charged and unpackaged* | 14 | 2 |
| *Confirmed participant is happy to continue being in the trial* | 12 | 4 |
| *Demonstrated how to assemble, fill, use and charge E-cigarette (EC) (using spare EC & mouthpiece covers)* | 15 | 1 |
| *Offered participants options to try different strengths and flavours* | 14 | 2 |
| *Provided EC Starter Kit (including charged EC unit, charger, plug) **WITHOUT** packaging* | 16 | 0 |
| *Provided Instructional (set-up) booklet/sheet and talked through* | 13 | 3 |
| *Provided E-cigarette support sheet and talked through* | 13 | 3 |
| *Supplied 5 bottles of preferred e-liquids (any combination)* | 16 | 0 |
| *Completed e-liquid log* | 16 | 0 |

As illustrated, the intervention was delivered with high fidelity across all regions and sizes of centre. In most cases, centre staff followed the intervention checklist that they had been provided with, including offering different flavours of e-liquids to participants, demonstrating how to fill up and use the e-cigarette device. Researchers noted that centre staff generally appeared confident when delivering the intervention and were encouraging and supportive, with good rapport between staff and participants. This was reflected in staff interviews:

> I: …how did you feel providing the initial instructions for the e-cigarettes?

> R: Yeah, fine, confident in doing so, wasn’t an issue. Obviously I had the sheet with me, so that was something that I would go off, and really it wasn’t a problem. (Female, Support Worker)

Most interventions took place in a private setting, generally a meeting room/office with a few being partially private, meaning that rooms contained windows looking into other areas. A smaller number were not private, either where the staff member was unable to leave the area unattended due to other staff cover not being available, or where space was limited and nowhere private was available.

Intervention delivery time was usually around 13 minutes (in line with expectations) with checklists only noting one occasion when the staff member cut a session short due to lack of time. Staff generally found time to deliver the intervention as arranged with the research team, however on a few occasions the staff member had to postpone due to other service user issues needing urgent attention.

Overall, fidelity to intervention checklists was high with no notable variations by region, size of centre or type of services delivered, suggesting that the intervention content had transferability across settings.

For follow up support sessions, fidelity to checklist components are summarised in Table 4:

**Table 4:** components of intervention support sessions.

| *Component* | *Discussed*<br>*(n)* | *Not*<br>*discussed*<br>*(n)* | *Missing*<br>*(n)* |
| --- | --- | --- | --- |
| *Checked they are happy to continue being in the trial?* | 7 | 1 | 8 |
| *Checked the participant still has the e-cigarette provided?* | 8 | 0 | 8 |
| *Checked whether the participant has used the e-cigarette provided?* | 8 | 0 | 8 |
| *Asked if they have experienced any burning and/or horrible taste when using the EC and if so suggest replacing (and help to replace the coil).* | 8 | 0 | 8 |
| *Checked whether the participant had any problems with the e-cigarette?* | 8 | 0 | 8 |
| *Supplied 5 bottles of preferred e-liquid* | 8 | 0 | 8 |
| *Completed e-liquid log* | 8 | 0 | 8 |

Follow-up observations were unable to be scheduled in several cases. Reasons included lack of availability of the intervention delivery staff and no available private spaces to meet with participants. Where follow-up did occur, fidelity to checklists was high however sessions were significantly shorter than initial sessions. Researchers noted that staff often seemed to be short of time and sessions felt rushed, with ongoing e-cigarette support impacted by staff shortages. Further still, at one site staff shortages meant a lack of communication about where e-liquids were being stored, impacting ability to re-distribute them to service users as planned.

### Implementation and any adaptations to the intervention

For most of the intervention sessions, delivery was one-to-one as intended. In one case, a friend of the participant attended to provide support and help them understand the content due to additional learning needs. In two other cases between two and four participants received the intervention together for convenience of staff involved.

There were examples of staff expanding on the checklist of the core intervention components to personalise or adapt delivery. In half of EC sites, it was noted that the staff member went beyond the core intervention components, expanding with other information they had learned in the training session. In some centres, staff drew on their own experiences as current users of e-cigarettes, having previously quit smoking and, in some cases, switching to e-cigarettes, in order to connect more with participants.

Participants were often allowed to try the EC in the centre during the intervention and in one case the staff member took the participant outside during the intervention delivery so they could test out the strengths and flavours.

Several staff interviewees involved in delivery reported wider scepticism among centre staff making delivery more challenging, most commonly among other staff not involved in the intervention than among service users:

> …there was nothing I was concerned about because I’d already done my own research, but…trying to get that message across to the people who I work for…I wish they’d been on that course…that part of it really did frustrate me, because I did that training courses. I took the information back… (Male, Support Worker)

> I: …so scepticism around e-cigarettes – would you say that was present, or not really, in the centre?

> R: Not really, not really. Not from the clients, anyway, more from the staff, I think it was, if anything, but not from the clients, no. (Male, Support Worker)

### Maintenance and evidence of sustained changes within centres

While sustained quit rates were not as high as estimated^16^ some staff noted that they knew of service users who had continued with smoking cessation efforts after the trial:

> I know of four clients and all four of them are getting on really, really well with it and they talk from a place of pride about how much they’ve cut down on their fags and they’ve actually gone out and bought themselves a different vape, a better one that’s more powerful. (Male, Support Worker)

However, other staff discussed barriers to embedding smoking cessation in continued centre life, citing practical concerns from a couple of staff who weren’t sure whether some participants would continue vaping once they were no longer supplied with e-liquids from the study:

> … well I just wonder if they’ll continue to use it now that the liquid is no longer available to them at the hostel. Now that they have to go and buy the liquid I’m not sure he would have the willpower to buy the liquid for his vape. (Female, Support Worker)

One staff member described sharing their personal stock of e-liquid with a service user to continue supporting them. Such practice of sharing personal supply is not feasible for longer-term support, suggesting that supply of e-liquids needs to be given for longer than was offered in the trial (4-weeks), and is likely important in promoting sustained change.

Most staff interviewees agreed that the intervention was aligned to the aims of their individual organisations (e.g., addressing health needs, promoting behaviour change), and believed the interventions added a new strand to the type of support they could offer:

> … I think it’s just more about being more aware of the dangers of smoking, and just generally giving those reminders to clients to let them know that, fair enough, we know you take these substances, and there are dangers to that, but smoking obviously has its dangers, well known, and they’re just as problematic as the Class A substances. So how about try and reduce it, and go to vaping, and seeing if you can reduce smoking completely. (Male, Support Worker)

Most staff felt that smoking cessation as a whole was a ‘fit’ with the existing harm reduction ethos underpinning their service delivery, suggesting potential for future embedding of cessation interventions with centres may be achievable:

> …we have the dental nurse in…we have the sexual health nurse in…I don’t see why…stopping smoking should be out of that equation the same as brushing your teeth…or your physical health as well… (Female, Support Worker)

There was also some evidence that a harm reduction narrative may also increase acceptability of the intervention being offered to the service users familiar with the existing harm reduction approaches:

> …a lot of our clients…are very aware of harm reduction, so it’s not a…foreign concept, and not a shameful thing…if you reduce a bit, then there’s loads of positives to gain from that, and I suppose we’re equally the environment that supports that… (Male, Manager)

However, reflecting the experience of researchers during centre recruitment, in some settings, it was suggested that smoking would be less of a priority than other, often more immediate, needs:

> …in terms of like hierarchy of harm, there would be…more pressing issues, I suppose, than smoking. Obviously, it’s very important as well, but…yeah, a bit different… (Female, Support Worker)

> …with somebody’s addiction issues, mental health issues, when they’re in crisis and they’ve not got a house…[smoking] wouldn’t be the first thing…I would prioritise their needs first…But if somebody wasn’t so chaotic…they were quite stable in their lifestyle…then, yes, we could say, oh…we have smoking cessation… (Female, Support Worker)

At times when centres are operating with financial constraints and challenges presented by staff turnover, dealing with those issues perceived as more urgent is likely to take precedence, suggesting that resourcing to support delivery of smoking cessation may be needed to embed the service within longer-term centre activities.

## Discussion

This process evaluation considered effective implementation of the e-cigarette intervention, in relation to fidelity and organisational or geographical differences. Findings suggest the feasibility of delivering this intervention in services supporting people experiencing homelessness, with little variation in relation to centre type, size and location. Implications for future work and recommendations are presented below.

### Feasibility and Fidelity of Implementation

The intervention was successfully delivered with high fidelity in participating centres, regardless of differences in size, geographic location, or centre type. This consistency suggests that the intervention is highly transferable across diverse settings with different challenges and across regions, (although transferability may be limited outside of Great Britain). As evidenced by feedback, the training provided to staff was well-received, equipping them with the knowledge and skills necessary to deliver the intervention effectively. Key intervention components, such as demonstrating e-cigarette usage and providing choices of nicotine strength and flavours, were implemented consistently, further supporting the intervention’s feasibility. Although staff at some centres did ask for the training to be shortened, this did not negatively impact on staff experience and should be considered for similar environments that may also require these types of training adaptions (e.g., prisons, mental health units). As well as providing practical knowledge on e-cigarettes and their use, the training was helpful in dispelling common misinformation on e-cigarettes. Future studies should consider delivery of training to all staff and participants within target settings (or as many as possible) to maximise benefits.

Nevertheless, some challenges in delivery were noted, including limited staff availability and time, in part due to the intervention largely being delivered by a limited number of the total staff who attended training. These issues occasionally impacted follow-up sessions, which were sometimes rushed or postponed due to competing demands.

Furthermore, while the intervention was designed to be integrated into routine centre activities, practical constraints, such as limited private spaces and time pressures, highlighted the importance of additional resourcing to support long-term delivery. These challenges and training needs were also highlighted in our feasibility study^7,15^.

### Acceptability and Contextual Fit

The intervention was largely acceptable within homelessness centres. Staff reported increased confidence in addressing smoking behaviours following training, as well as feeling more able to challenge inaccuracies about e-cigarettes when stated by both service users and other colleagues. However, some scepticism among non-participating staff, as well as some cases of managerial resistance to vaping policies in certain centres underscored the need for broader training and intervention buy-in.

The intervention’s alignment with the harm reduction ethos prevalent in homelessness services was a notable strength. Staff emphasised that tobacco harm reduction approaches complemented existing health and wellbeing initiatives, such as sexual health and dental care services. However, the complex needs often evident in this population group meant that centres sometimes needed to addressed immediate and acute needs, such as housing or other substance support, over smoking. This, coupled with identified pressures on staff time and capacity, may suggest that, for long-term sustainability a dedicated stop smoking advisor situated in centres, is best placed to deliver smoking interventions, reducing pressure on staff but still maintaining support for service users.

### Sustained Impact and Challenges

As reported elsewhere^23^ evidence of sustained behaviour change among service users was mixed. While some participants continued using e-cigarettes after the study, others faced barriers such as the cost of e-liquids once trial-provided supplies were exhausted. Staff interviews suggested that ongoing provision of resources, such as e-liquid supplies, may be critical for supporting longer-term use. Additionally, embedding smoking cessation as a routine aspect of centre services would require addressing structural barriers, including staff shortages and funding constraints.

### Implications for Future Delivery

Findings suggest several recommendations to enhance future intervention implementation and sustainability. First, expanding training to include all staff, not just those directly delivering the trial intervention, could foster a more supportive environment and address scepticism. Where possible, providing information aimed at challenging e-cigarette misinformation among service users should also be considered. Second, securing resources for sustained provision of e-cigarettes and related supplies would mitigate financial barriers for service users. Third, integrating smoking cessation interventions into broader health promotion strategies within centres may improve prioritization. This includes exploration of greater alignment and potential co-delivery opportunities with existing services. Finally, addressing systemic challenges, such as high staff turnover and resource limitations, will be critical for scaling up and embedding such interventions in homelessness support services.

## Strengths and limitations

This study provides novel insights into the delivery of trials with an overlooked population that can be of value to researchers and policy makers. Centre and participant recruitment and retention shows signs of promise that both the intervention and data collection were acceptable to staff and support centres. However, there are limitations which should be acknowledged. While retention was higher than targets set out in the study protocol, there was significant attrition between baseline and 24 weeks and the reasons for this are unable to be explored, meaning we cannot be sure that those who were not retained did not differ in significant ways. There may also have been potential biases in qualitative data, as interviewees may have been more engaged or positively inclined toward the intervention. Additionally, while the sample represented diverse centre types, findings may not be generalizable to all homelessness support settings in the UK or in countries outside of the UK.

## Conclusion

The process evaluation of the SCeTCH trial indicates effective implementation of the intervention, with high fidelity and little organisational or geographical differences. It also demonstrates the acceptability of delivering a smoking cessation intervention in homelessness services, with positive staff and service user feedback highlighting the intervention’s potential to address smoking-related health inequalities among people experiencing homelessness. The findings provide valuable insights into the facilitators and barriers to implementation, with implications for future smoking cessation efforts in similar contexts, which is becoming increasingly important as smoking rates decrease in the general population but remain higher in certain subpopulations. Addressing practical and systemic challenges will be essential for scaling up and sustaining such interventions in the future as part of the UK’s ambition to be ‘smokefree’ by 2030^25^.

## Data Availability

All data produced in the present study are available upon reasonable request to the authors

## Author contributions

### CREDIT statement

Rachel Brown (*Funding acquisition, Methodology, Supervision, Writing – Original Draft)*, Allison Ford (*Funding acquisition, Methodology, Supervision, Formal Analysis, Writing – Reviewing and Editing)*, Laura Reid (*Writing – Reviewing and Editing)*, Lauren McMillan *(Investigation, Formal Analysis, Writing – Reviewing and Editing)*, Kirstie Soar (*Data Curation, Investigation, Formal Analysis, Project Administration*, *Writing – Reviewing and Editing)*, Francesca Pesola (*Data Curation, Formal Analysis*, Validation, *Writing – Reviewing and Editing*, Caitlin Notley (*Funding acquisition, Methodology, Supervision, Writing – Reviewing and Editing)*, Emma Ward *(Investigation, Formal Analysis)*, Bethany Gardner *(Investigation, Formal Analysis)*, Anna Varley *(Investigation, Formal Analysis)*, Charlotte Mair *(Investigation, Formal Analysis)*, Jessica Lennon *(Investigation, Formal Analysis)*, Janine Brierley *(Investigation, Formal Analysis)*, Amy Edwards *(Investigation)*, Danielle Mitchell *(Investigation)*, Debbie Robson *(Resources, Investigation),* Peter Hajek *(Methodology, Data Curation, Formal Analysis),* Allan Tyler *(Investigation, Methodology, Data Curation, Formal Analysis)*, Steve Parrott, *(Methodology, Data Curation, Formal Analysis),* Jinshuo Li *(Methodology, Data Curation, Formal Analysis),* Linda Bauld *(Supervision, Validation)*, Sharon Cox *(Conceptualisation, Funding acquisition, Methodology, Supervision, Writing – Reviewing and Editing)*.

### Declaration of generative AI and AI-assisted technologies in the writing process

During the preparation of this work the author(s) used no AI or AI assisted technologies.

## Supplementary files

**Supplement 1: Details of SCeTCH, a cluster randomised control trial** (reproduced from: Ford et al. 2025^21^)

### Design

A multi-centre two-arm cluster randomised controlled trial (see Cox et al. 2022 for full protocol).

### Intervention

Participants in the EC arm were provided with a tank-style refillable EC (PockeX), a four-week supply of e-liquid with a choice of nicotine strength (12mg/mL and 18mg/mL) and flavours (tobacco, menthol, or fruit), and a brief introductory session on how to use the EC, and an EC fact sheet.

Participants in the usual care (UC) arm were provided with Very Brief Advice PLUS (VBA+) on smoking, a quit smoking support leaflet and signposting to the local Stop Smoking Service (SSS), including help to facilitate referral if required.

The EC intervention and UC were delivered by centre staff following staff training from the research team.

### Participants

Thirty-two homelessness support centres (clusters) from across Great Britain were recruited to the study. Sixteen centres were randomised to EC (n=239 participants) and 16 to UC (n=238 participants).

Participants were aged 18+, self-reported smoking daily (verified by centre staff), regularly attended one of the centres, and were not currently using a smoking cessation aid. Participants did not have to be motivated to quit and there was no agreement that a cessation attempt should be made.

**Study outcomes** (reported in full Dawkins et al. 2024)

The primary trial outcome was sustained carbon monoxide (CO) validated smoking cessation at 24 weeks. There was no difference in sustained 24-week smoking cessation rates between the EC arm vs. the UC arm (aRR:2.43, 95%CI: 0.51-11.64)

The secondary trial outcomes were i) 50% smoking reduction at 24 weeks, ii) 7-day point prevalence quit rates at 4, 12 and 24 weeks, and iii) self-reported changes in risky smoking practices from baseline to 4,12 and 24 weeks. i) A higher number of participants in the EC arm reduced their smoking by 50% at 24 weeks (aRR:2.02, 95%CI:1.44-2.84) compared to those in the UC arm. ii) A higher number of participants self-reported and had CO-validated 7-day point prevalence abstinence at 24 weeks (aRR:2.95, 95%CI:1.05-8.29), and self-reported 7-point prevalence abstinence at 4 weeks (aRR:3.32, 95%CI:1.34-8.23) compared to UC. iii) There was no main effect of the intervention on the reduction of sharing cigarettes (aOR=1.03, 95%CI: 0.38-2.77), smoking a discarded cigarette (aOR=0.28, 95%CI: 0.07-1.06), or asking strangers for cigarettes (aOR=1.61, 95%CI: 0.55-4.77).

